# A unified meta-regression model identifies genes associated with epilepsy

**DOI:** 10.1101/2024.06.27.24309590

**Authors:** Oscar Aguilar, Mijail Rivas, Manuel A. Rivas

**Affiliations:** Department of Management Science & Engineering, Stanford University; National Institute of Neurology and Neurosurgery, Mexico and Clinical Epileptology Fellowship UNAM; Department of Biomedical Data Science, Stanford University

## Abstract

Epilepsy is a highly heterogeneous disorder thought to have strong genetic components. However, identifying these risk factors using whole-exome sequencing studies requires very large sample sizes and good signal-to-noise ratio in order to assess the association between rare variants in any given gene and disease. We present an approach for predicting constraint in the human genome through application of a Hidden Markov Model (HMM) to whole exome sequencing (WES) data. Using the Regeneron Genetics Center Million Exome dataset and the AllofUs whole genome sequencing data, we predict the probability of observing no variants across the population for each position in the genome. We then incorporate the predictions with the “rejected substitutions” (RS) score from Genomic Evolutionary Rate Profiling (GERP), pathogenicity predictions from AlphaMissense (AM), and pLoF/Missense annotations from Epi25 into a model that detects epilepsy-associated genes. We identify a set of significant (*p* < 3. 4 × 10^−7^) genes which did not meet exome-wide significance in previous studies: *KCNQ2, SCN2A, STXBP1, CACNA1A, SLC6A1, DYRK1A, KCNB1, SATB1, PCDHAC2, SP4*, and *RYR2*,. Our models allow us to evaluate the contribution of constraint, protein structure based pathogenicity prediction from AM, and pLoFs jointly. We show that unifying these moderators into a single model allows us to both strengthen our evidence for genes with already-known links to epilepsy and also identify new genes with likely links to epilepsy.

## Introduction

Epilepsy is one of the most common neurological disorders, affecting individuals across all ages and populations.^1^ It encompasses a diverse spectrum of phenotypes and is marked by a predisposition to recurrent epileptic seizures, which can lead to substantial cognitive, psychological, and social consequences.^2^ Beyond its clinical impact, epilepsy represents a major global health burden, affecting approximately 70 million people worldwide, and the cost of diagnosis and treatment exceeds USD 100 billion annually.^2^ While epilepsy can result from various causes such as infection, trauma, and stroke, advances in genomics have demonstrated that genetic factors are likely implicated in over two-thirds of cases where the cause is unknown.^3^

Advances in sequencing and gene discovery have enabled the study of the genetic etiologies of epilepsy.^4–6^ Genetic inheritance can be polygenic, as seen in genetic (idiopathic) generalized epilepsy (GGE [MIM: 600669]), where pathogenic variants are usually not detected in standard gene panels. Alternatively, the inheritance can be monogenic, as observed in early-onset developmental epileptic encephalopathies (DEE [MIM: 308350]), with pathogenic variants identified through epilepsy gene panels or whole-exome sequencing. These insights have motivated the integration of genetic testing into clinical care, with benefits including refined diagnosis, targeted therapy, and informed genetic counseling.^7^ Despite these advances, the delineation of genes that confer risk for epilepsy remains incomplete. There is no consensus definition of what constitutes an “epilepsy gene,” and distinctions are often made between genes directly implicated in epilepsy pathogenesis and genes associated with broader syndromic presentations that include seizures.^8–11^

Whole-exome sequencing (WES) has emerged as a powerful tool for investigating the genetic basis of epilepsy, enabling the identification of rare variants associated with various diseases. A landmark study by the Epi25 Collaborative analyzed over 54,000 individuals (20,979 epilepsy cases and 33,444 controls), yielding novel insights into the genetic basis of epilepsy across multiple subtypes and ancestries.^6^ However, the study also highlighted key challenges: the rarity and heterogeneity of causal variants, limited statistical power for gene-level association testing, and the need for improved variant interpretation frameworks.

A key strategy for improving rare variant association studies involves incorporating prior knowledge about *constraint*, defined *as* the evolutionary intolerance of a region to variation. Existing approaches such as the probability of loss-of-function intolerance (pLI) and the Loss-of-function Observed/Expected Upper bound Fraction (LOEUF) from the gnomAD consortium quantify gene-level depletion of protein-truncating variation relative to expectation.^12^ Other regional metrics such as the Missense Tolerance Ratio (MTR) evaluate local intolerance to missense variation.^13^ Likewise, evolutionary conservation-based measures such as Genomic Evolutionary Rate Profiling (GERP) estimate constraint from multispecies alignments, providing position-specific “rejected substitution” (RS) scores that quantify the deficit of substitutions relative to neutrality.^14–16^ GERP captures constraint at single-base resolution and complements population-based depletion metrics by emphasizing evolutionary conservation across mammals. Collectively, these constraint scores have become essential tools for gene discovery and variant prioritization, yet they are typically static, population-derived summaries that do not fully capture the spatial structure or context of mutational depletion.

Recent advances in deep learning have further improved variant-level prioritization. AlphaMissense, a state-of-the-art model developed by Google DeepMind, predicts the pathogenicity of all possible missense variants using protein structure-informed features derived from AlphaFold2.^17^ This resource provides a genome-wide atlas of predicted missense effects, and complements traditional constraint metrics by leveraging structural and functional priors.

To address the challenge of limited statistical power in rare variant association studies, we developed an approach that leverages sequence-level constraint inference using a Hidden Markov Model (HMM). We applied this framework to large-scale exome data from the Regeneron Genetics Center (RGC-ME)^13^ and validated its robustness using the All of Us cohort.^18^ By integrating this sequence-derived constraint signal with evolutionary constraint scores from GERP, structure-informed pathogenicity predictions from AlphaMissense, and annotations of predicted loss-of-function (pLoF) and missense variants, we construct a unified meta-regression statistical model to assess gene-level associations with epilepsy risk. This integrative method enables the detection of biologically meaningful signals that are missed by standard approaches, particularly in the context of heterogeneous neurodevelopmental disorders like epilepsy.

## Methods

### Inferring Constraint in the Human Genome Using a Hidden Markov Model

To build on prior gene- and region-level constraint metrics and improve resolution at the nucleotide level, we develop a Hidden Markov Model (HMM) to infer constraint directly from large-scale human variation data. An HMM is a statistical framework that models a sequence as a series of underlying, unobserved states—in this case, mutationally constrained or tolerant positions—that generates the observed genetic variation. By learning how likely the sequence transitions between these hidden states, the model can infer which regions of the genome are under selective constraint even when direct evidence is sparse. The HMM is particularly well-suited to this task due to its ability to model spatial dependencies in sequence data, capturing the transition between mutationally tolerant and constrained regions more effectively than simpler frequency-based approaches.

We define the observed sequence for each chromosome as a binary vector, where each position is encoded as “1” if at least one variant is detected in the cohort and “0” otherwise. The latent states represent the underlying mutational constraint: one state corresponds to constrained regions, where mutations are rare due to selective pressure, while the other represents non-constrained regions. The HMM is trained using the expectation-maximizing Baum-Welch algorithm, which iteratively updates the transition and emission probabilities to maximize the likelihood of the observed data.

Our implementation uses the `hmmlearn` library in Python to simulate a genomic sequence of specified length with a biased probability of mutation to reflect realistic variation rates. Before training, the observed binary sequence is preprocessed to account for local dependencies between adjacent sites by summarizing short subsequences of length “order.” These subsequences are converted into count-based representations, in which occurrences of 0s and 1s are aggregated to support more stable statistical estimation. This preprocessing step enables the model to better capture the short-range structure of mutational patterns, improving its ability to distinguish between constrained and tolerant regions.

Using these generated sequences, we initialize and fit a first-order HMM with two hidden states, representing constrained and non-constrained genomic regions. The model is trained using the Baum–Welch algorithm, which iteratively estimates how likely each genomic position is to belong to either the constrained or unconstrained state and updates the underlying probability of transition between these two states. Conceptually, the algorithm alternates between two steps: estimating which regions are most likely constrained given the current model (the *expectation* or E-step) and then refining the model parameters to better explain the observed mutation patterns (the *maximization* or M-step).

Formally, the expectation (E) step computes the *forward* and *backward* probabilities for each position *t* along the observed sequence. The *forward probability*, denoted α_t_ (*i*), represents the joint probability of observing the sequence up to position *t* and being in hidden state *i* at that position. Conversely, the *backward probability*, β_*t*_ (*i*), represents the probability of observing the remaining sequence from position *t* + 1 to the end, given that the model is currently in state *i* at position *t*. Using these quantities, we compute (1) the posterior probability of being in state *i* at position *t*, γ_*t*_ (*i*), and (2) the joint posterior probability of transitioning from state *i* to state *j* between positions *t* and *t* + 1, ξ_*t*_ (*i, j*). In the maximization (M) step, the parameters of the model are updated to maximize the expected complete-data log-likelihood. This iterative process continues until convergence, yielding the most probable configuration of constrained and tolerant segments across the simulated genome.

To ensure robustness, we exclude positions with insufficient sequencing depth, retaining only those with >20% coverage across at least 90% of individuals. This criterion mitigates biases introduced by missing data and ensures that the inferred constraints reflect true biological patterns rather than sequencing artifacts. Additionally, we restrict our analysis to protein-coding regions, as these sites are most relevant for disease association studies. Once trained on chromosome 2 from the RGC-ME dataset, we use the model to infer constraint probabilities across all chromosomes.

### Validation of Constraint Predictions Using an Independent Dataset

To validate the robustness and generalizability of our constraint inference model, we apply it to an independent whole-exome sequencing dataset from the AllofUs (AoU) initiative, comprising 245,388 individuals of mixed ancestry. This step is crucial in demonstrating that the HMM captures fundamental biological properties rather than dataset-specific artifacts.

We use the trained HMM from chromosome 2 of RGC-ME to predict constraint probabilities across all other chromosomes in AoU to maintain independence from the training set. To assess the validity of our predictions, we examine the joint distribution of constraint probabilities across both datasets in Figure 2a. If our model captures meaningful constraint information, we expect consistent predictions between RGC-ME and AoU, especially for highly constrained and highly mutable regions.

**Figure 1:**
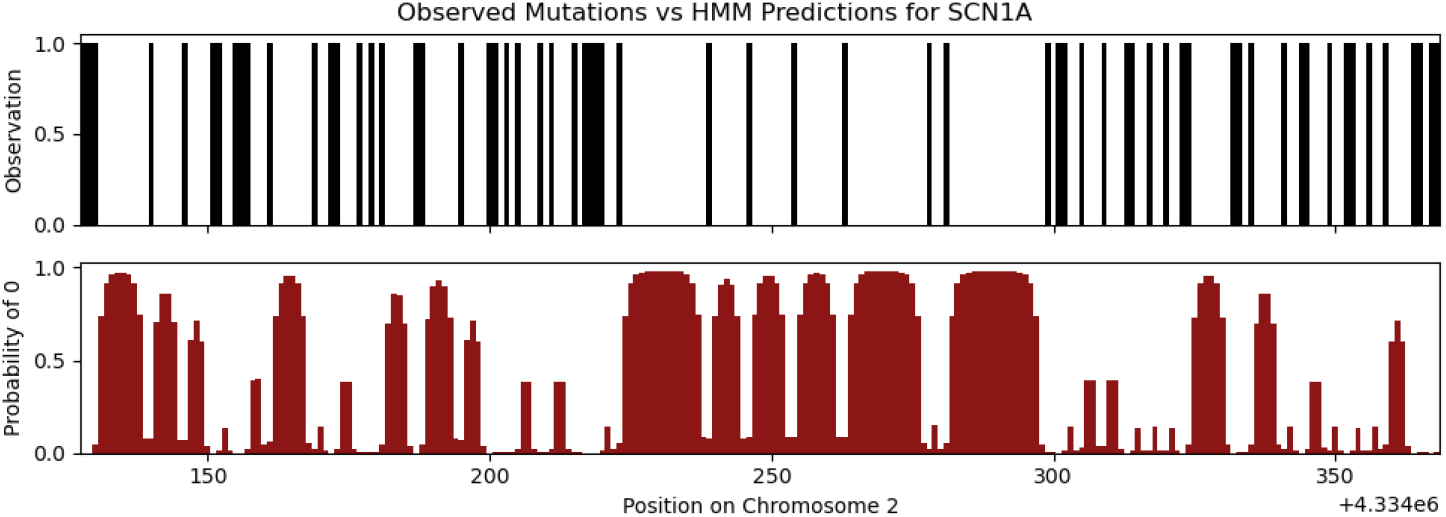
Example of an observed binary sequence vs the predictions made by the HMM.

**Figure 2:**
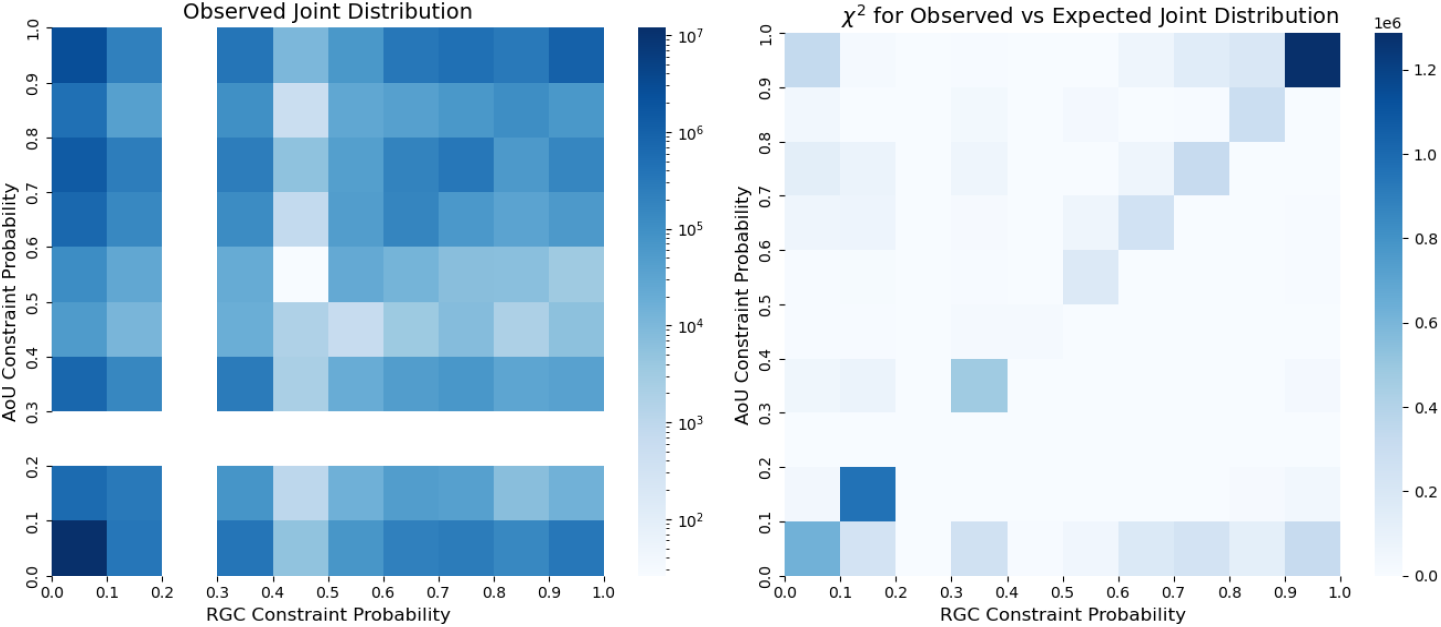
Observed joint distribution (left) of WES constraint predictions (excluding chromosome 2) from AoU and RGC, and distribution of corresponding *χ*^2^ statistics (right). The [0.2, 0.3) bin has no observations because the HMM does not generate constraint values within that range.

Our analysis confirms a strong overlap in constraint predictions between the two datasets, demonstrating the model’s reliability and consistency across cohorts. The highest density of observed probability mass is concentrated in regions with low predicted constraint (< 0.1), consistent with the expectation that most genomic positions are not under strong selective pressure. Importantly, high-probability constraint predictions (top decile) show significant concordance between RGC-ME and AoU, indicating that the HMM identifies constrained regions independently of the dataset on which it was trained. To quantify this relationship, we compare in Figure 2b the observed joint distribution to the expected distribution under an independence assumption. The resulting chi-square statistics reveal significant deviation from independence in both low- and high-constraint probability regions, with values ranging from 10,000 to 90,000 across chromosomes. In contrast, regions where the model disagrees (i.e. infers high constraint in one dataset and low constraint from the other) exhibit lower chi-square values (3,000–37,000), suggesting that discordant predictions are limited and largely confined to intermediate-probability regions. Together, these results support that our model captures stable and biologically meaningful patterns of constraint that generalize across datasets, populations, and genomic contexts.

### Comparison with Existing Constraint Metrics

To evaluate the HMM predictions as a measure of mutational constraint, we compare them to three widely used gene constraint metrics: the Missense Tolerance Ratio (MTR), the missense z-score, and the Genomic Evolutionary Rate Profiling (GERP). The MTR, derived from the gnomAD dataset, is defined as the observed proportion of missense variants divided by the expected proportion, relative to the number of all possible variants in a given genomic window.^13^ The missense z-score, on the other hand, quantifies deviation from expectation by computing the z-score of the observed missense variant count relative to the expected count, as derived from the gnomAD dataset.^12^ A higher missense z-score indicates stronger selective constraint, meaning fewer missense variants were observed than expected, whereas a lower MTR value suggests higher intolerance to variation. GERP, in contrast, measures evolutionary constraint across 35 mammalian species by estimating the deficit of substitutions relative to neutrality. Positive RS scores indicate purifying selection and evolutionary conservation, providing a cross-species complement to population-based metrics such as MTR and the missense z-score.

To ensure a fair comparison between our HMM predictions and these established constraint metrics, we aggregate our model’s constraint probabilities at the gene level by computing the proportion of each gene where the HMM assigns a probability greater than 0.5 to the constrained state. This aggregated “constraint proportion” is then analyzed in relation to the MTR, missense z-score, and GERP score. Figures 3a and 3b illustrate the joint distributions of predicted constraint proportions versus the first two measures, showing broadly consistent trends: genes predicted to be highly constrained by the HMM tend to have lower MTR values and higher missense z-scores. The relationships are quantified by fitting a generalized linear model (GLM) using ordinary least squares regression, yielding R-squared values of 0.204 and 0.146, respectively. These results confirm that the HMM captures gene-level constraint patterns comparable to those observed in established population-based metrics, while offering improved spatial resolution and interpretability.

**Figure 3:**
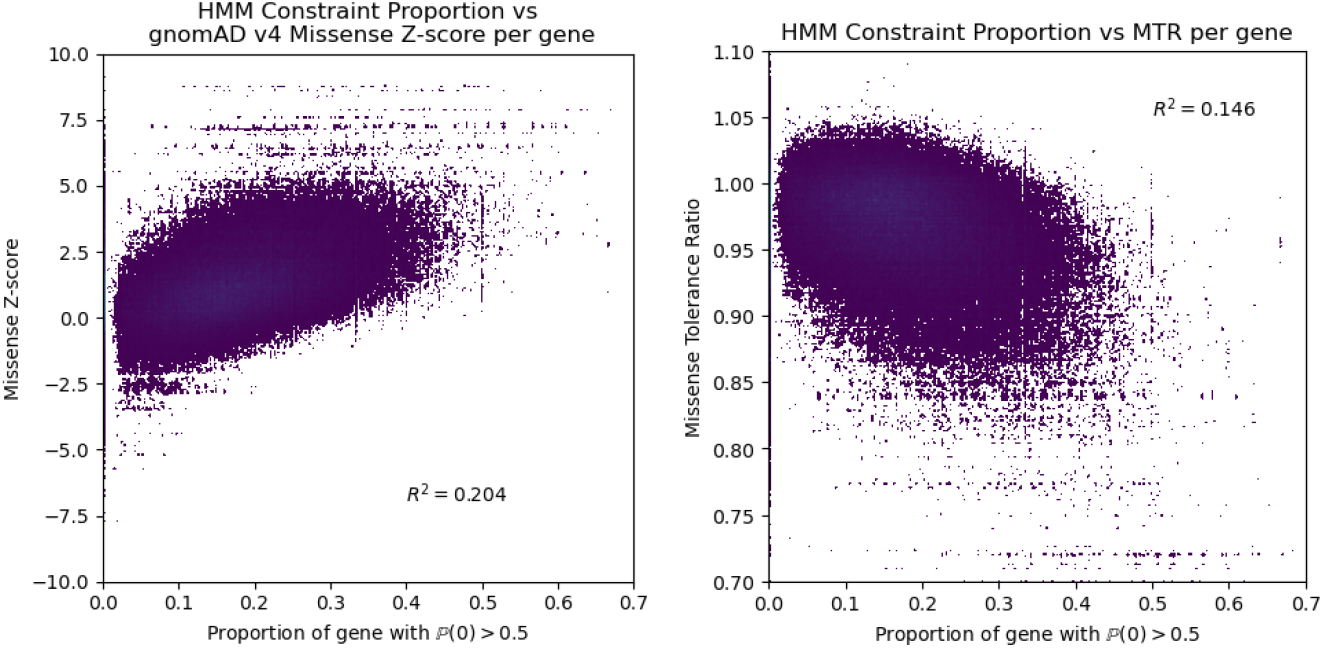
Joint distribution of z-score of observed vs expected missense variants per gene (left) and missense tolerance ratio (right) versus proportion of gene with HMM constraint probability > 0.5 (x-axis).

We next compare our HMM-derived nucleotide-level constraint predictions with GERP RS scores, which quantify evolutionary conservation at single-base resolution. Unlike the MTR and missense z-score, which are computed per gene, both the HMM and GERP operate at the locus level, allowing direct exome-wide comparison of their joint distributions. Across all positions, the correlation between HMM constraint probabilities and GERP RS scores is weak (*R*^2^ = 0. 006), indicating that the two approaches capture distinct aspects of constraint. However, analysis of their observed versus expected joint distribution in Figure 4 reveals a pronounced deviation from independence. Highly conserved sites (GERP > 4–5) that are not classified as constrained by the HMM are over-represented, whereas HMM-constrained but evolutionarily unconserved sites are under-represented. This bifurcation suggests that while GERP primarily reflects deep evolutionary conservation across mammals, the HMM captures more recent or population-specific selective pressures. Together, these complementary signals highlight that the HMM generalizes constraint detection beyond static evolutionary conservation, capturing mutational intolerance shaped by both functional and demographic processes.

**Figure 4:**
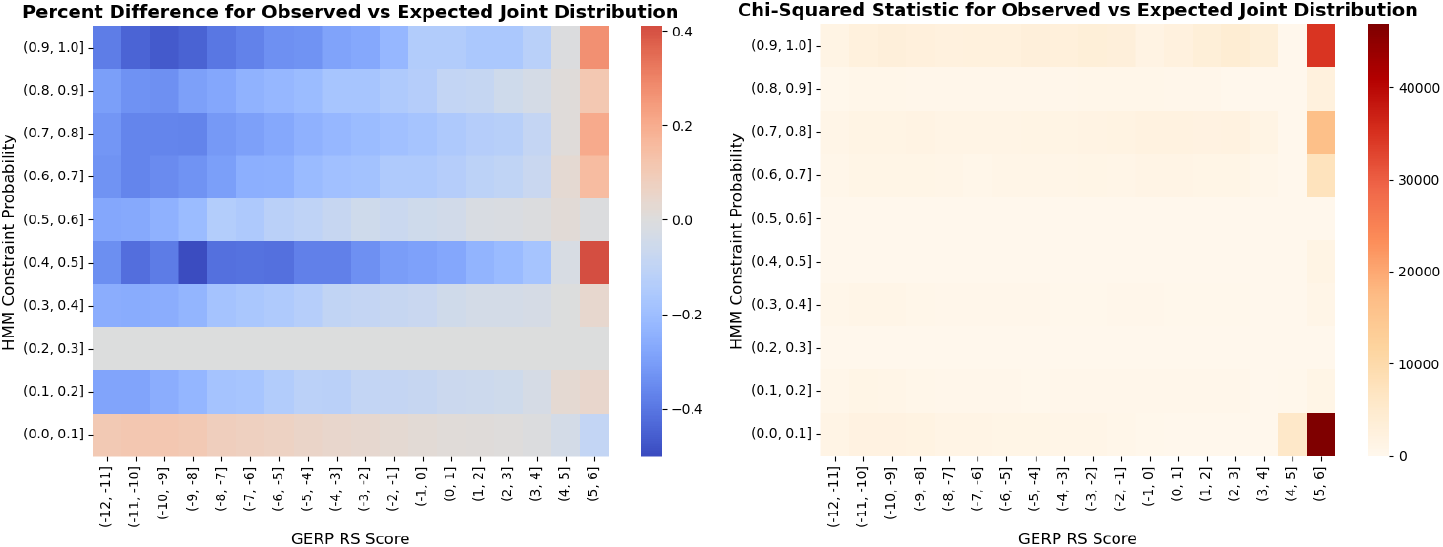
Distribution of percent difference (left) and *χ*^2^ statistics (right) for the observed vs expected joint distribution of HMM constraint predictions and GERP RS scores.

By comparing our model’s predictions to established population-based and evolutionary measures of selective constraint, we demonstrate that the HMM captures known signatures of mutational intolerance while also providing complementary information beyond existing metrics. The observed correlations with MTR and missense z-scores confirm that the HMM aligns with established gene-level patterns of constraint, whereas its partial independence from GERP indicates that it captures additional signals shaped by population-level variation and functional context rather than deep evolutionary conservation. Together, these findings support the HMM as a robust and biologically informative predictor of constraint, offering improved resolution and interpretability for identifying functionally important regions of the genome.

Lastly, we assess the marginal informativeness of constraint versus pathogenicity scores using logistic regression. Modeling observed variant presence as a function of AM pathogenicity scores alone yields a McFadden pseudo-*R*^2^ of 0.0081. In contrast, incorporating HMM-based constraint probabilities as predictors results in a much higher pseudo-*R*^2^ of 0.3834, underscoring the biological distinctiveness and predictive value of the HMM-inferred constraint signal over purely sequence-derived pathogenicity scores.

### Unified meta-regression model

To improve detection of genes associated with epilepsy, we integrate our HMM-derived constraint predictions with the “rejected substitutions” (RS) score from Genomic Evolutionary Rate Profiling (GERP), missense variant pathogenicity probabilities from AlphaMissense (AM) and loss-of-function (pLoF) and missense annotations within a unified statistical framework. We present a weighted least squares (WLS) meta-regression model to effectively weight each individual moderator (HMM, GERP, AM, pLoF, and missense) to minimize prediction variance and maximize accuracy, which was developed concurrently with Lauer and Rivas.^19^

Suppose that for gene *k*, we have variants *i* = 1, 2, …, *n*_*k*_. Then, for each moderator, we obtain the following outputs:

- 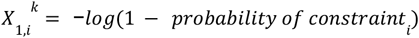, from the Hidden Markov Model (HMM);
- 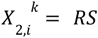, “rejected substitutions” (RS) score from GERP;
- 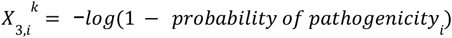, from AlphaMissense;
- 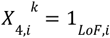 an indicator of whether variant *i* is a pLoF variant.
- 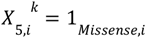 an indicator of whether variant *i* is a Missense variant.

The column vector 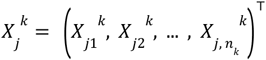 denotes the output from moderator *j* for all variants of gene *k*. Similarly, we define the *n*_*k*_ × 5 matrix 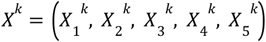.

For variant *i* of gene *k*, we have four distinct populations: (1) the subset of the population with variant *i* that is (1) epileptic (*ALT*_*EP*_), and (2) *not* epileptic (*ALT*_*CON*_), and the subset of the population without variant *i* that is (3) epileptic (*REF*_*EP*_), and (4) *not* epileptic (*REF* _*CON*_).

Let 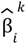 denote the estimated single-variant effect size estimate representing the log odds ratio of finding variant *i* of gene *k* in the population of patients with epilepsy versus the control. Using the following formula from the National Institute of Standards and Technology (NIST), ^20^ we obtain

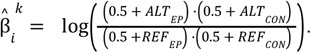

We can then estimate the standard error 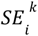 of estimated single-variant effect size 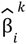, computed by

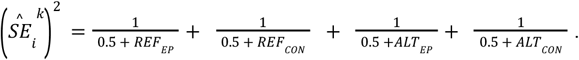

Each variant contributes a datapoint to the model; thus, we can weigh each of the *n*_*k*_ variants by positive weight 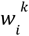 to achieve an aggregate effect size 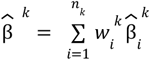. To minimize the variance of this aggregated estimator, we take 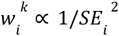 for all variants *i* of gene *k*. Let 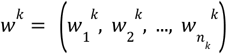 and *W* = *diag*(*w*^*k*^). Then, for any gene *k* and any variant *i* ∈ {1, 2, …, *n* _*k*_}, the WLS meta-regression model is defined as

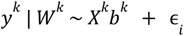

where *b* ^*k*^ is a vector of coefficients given by *b*^*k*^ = ((*X* ^*k*^)^⊤^ *W*^*k*^ *X*^*k*^)^−1^(*X*^*k*^)^⊤^ *W*^*k*^ *y* ^*k*^, and *y*^*k*^ is the vector of predicted effect sizes estimating 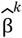.

To evaluate the relative explanatory power of each moderator, we compute the proportion of variance explained by each predictor using partial 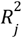 values:

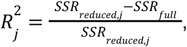

where *SSR*_*full*_ is the residual sum of squares from the full model, and *SSR*_*reduced*,*j*_ is the residual sum of squares from the model excluding moderator*j*. The total model *R*^2^ is:

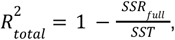

with 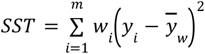 and 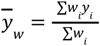.

We further test the significance of each predictor using the *t*-statistic:

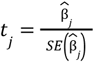

and assess overall model fit using the F-statistic:

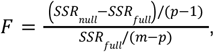

with corresponding unified model p-value *p*_*unified*_ = *P*(*F* > *F*_*observed*_).

Prior to fitting the meta-regression, we applied variant- and gene-level inclusion filters to ensure model stability and interpretability. Specifically, we retained only exonic variant positions with at least one observed alternate allele in the study population (cases or controls) and no more than five total alternate-allele counts across the combined cohort, thereby restricting analysis to rare, observed variants. Furthermore, we required that each gene–group meta-regression include at least N > 25 variants, following recommendations by Jenkins and Quintana-Ascencio.^21^ This threshold minimizes overfitting and ensures that estimated regression coefficients and partial *R*^2^ values are derived from adequately sized variant sets.

Variants are mapped to genes using a standardized gene identifier (Ensembl gene ID) joined to an HGNC gene symbol via our gene reference table. We restrict to autosomal exonic variants (X, Y, MT excluded) with non-missing consequence annotations. For Epi25, we retain variants whose consequence is either pLoF or missense after excluding synonymous, non-coding, and undefined annotations. We encode two binary indicators using the annotations:

- *consequence* ∈ {*pLoF*} → 1 _*LoF*,*i*_
- *consequence* ∈ {*damaging missense, other missense*} → 1 _*Missense*,*i*_

For SCHEMA, we exclude non-coding/synonymous classes: 5′/3′UTR, intronic, intergenic, upstream/downstream, non-coding transcript (exon), splice-region, mature miRNA, coding sequence, synonymous, and undefined annotations. We encode the binary indicators using:

- {*stop gained, splice acceptor, splice donor, frameshift*} → 1 _*LoF*,*i*_
- {*inframe insertion*/*deletion, stop retained*/*lost, protein altering, missense MPC*} → 1 _*Missense*,*i*_
- These dataset-specific consequence classes are used to construct the moderator indicators 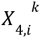 and 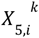 for the meta-regression model.

## Results

Having validated the HMM as a sequence-dependent predictor of mutational constraint, we next integrate its output with structure-based missense variant pathogenicity predictions from AlphaMissense, existing annotations of predicted loss-of-function (pLoF) variants, as well as GERP RS scores. These signals are combined within the weighted meta-regression framework described in the previous section to jointly estimate variant-level effects.This unified model provides a general framework for variant-level prioritization and gene-level inference of rare variant burden across diverse disorders. We first evaluate its performance in epilepsy, where extensive sequencing resources enable systematic benchmarking, and then demonstrate its broader applicability using a schizophrenia dataset with more limited power. Together, these analyses illustrate the model’s phenotype-agnostic design, in which constraint, pathogenicity, and pLoF information are flexibly integrated to detect disease-relevant genetic signals.

### Epilepsy

We apply our analysis to the Epi25 Collaborative dataset comprising 54,000 individuals with and without epilepsy.^6^ Following this study, we define exome-wide significance as *p* = 3. 4 × 10^−7^, corresponding to a Bonferroni correction across 18,531 protein-coding genes tested for four epilepsy subtypes and two variant classes. Genes with *p* < 1. 0 × 10^−4^ are considered suggestive. Table 1 compares the p-values for each moderator as well as our unified model for a subset of either (a) genes above the exome-wide significance threshold on our unified model and below the threshold in the Epi25 study or (b) suggestive genes with either known links to epilepsy or other neurological functions whose association with epilepsy were previously inconclusive. The genes meeting criterion (b) are designated by the pink-highlighted rows in Table 1. Table S1 in Supplementary Materials contains a full set of suggestive genes, and all results can be explored in our epilepsy dashboard (https://epilepsy-dashboard.streamlit.app/) Across the genes in Table 1, the meta-regression identifies statistically significant signals where prior population-based analyses lacked sufficient power for detection. In particular, the weighted meta-regression model is able to leverage the strengths of each individual moderator in the regression model to exact a stronger signal on several genes’ influence on the presentation of epilepsy.

**Table 1:**
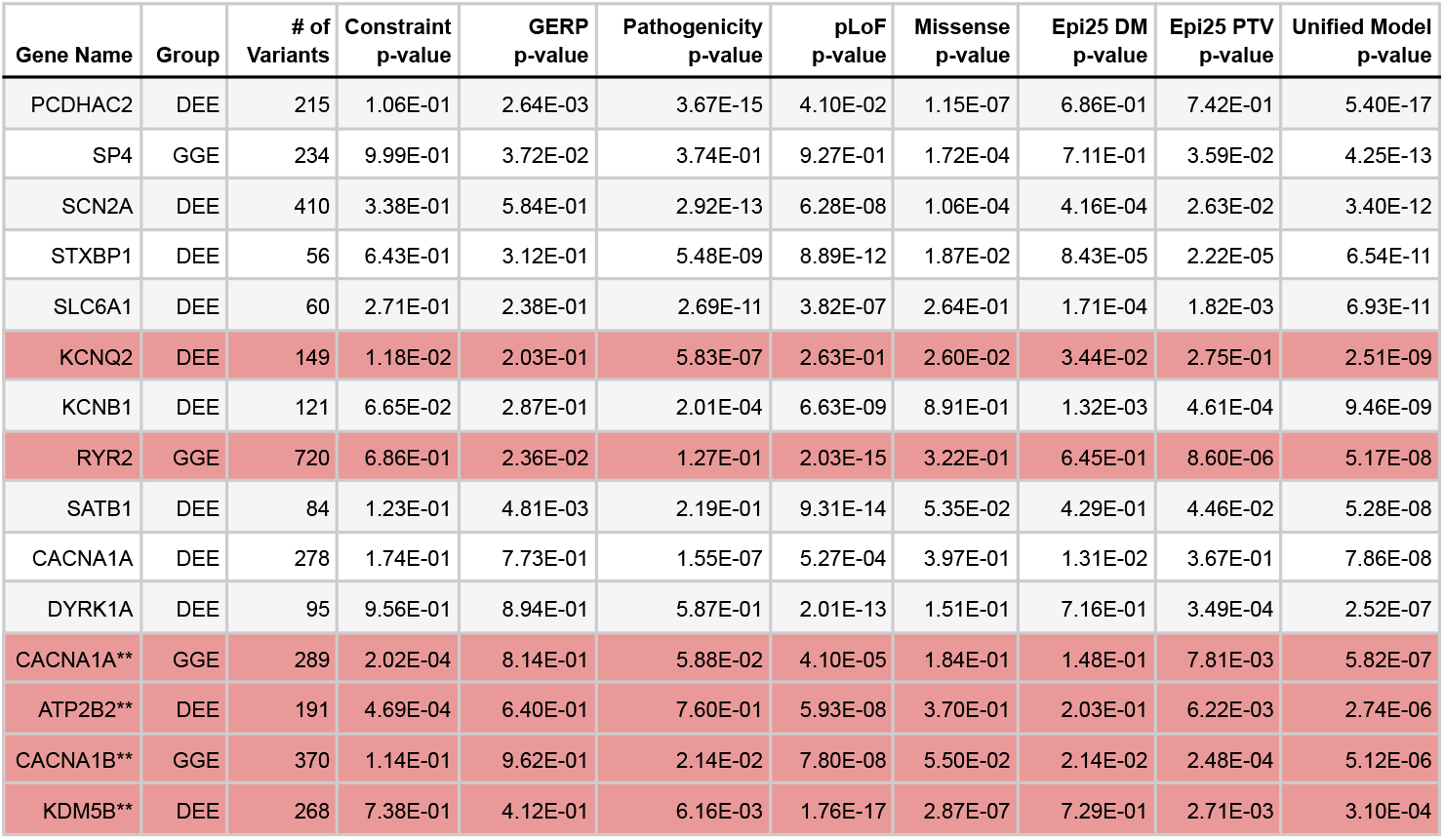
Comparison of p-values for each moderator of the unified model, the unified model value, and the Epi25 damaging missense and PTV p-values across four epilepsy gene groups. Results are the set of significant (*p* < 3. 4 × 10^−7^) gene-group pairs which did not meet exome-wide significance in the Epi25 study, as well as four important gene-group pairs not quite meeting this threshold, denoted by asterisks. The highlighted rows denote genes with known neurological function.

| Gene Name | Group | # of Variants | Constraint p-value | GERP p-value | Pathogenicity p-value | pLoF p-value | Missense p-value | Epi25 DM p-value | Epi25 PTV p-value | Unified Model p-value |
| --- | --- | --- | --- | --- | --- | --- | --- | --- | --- | --- |
| PCDHAC2 | DEE | 215 | 1.06E-01 | 2.64E-03 | 3.67E-15 | 4.10E-02 | 1.15E-07 | 6.86E-01 | 7.42E-01 | 5.40E-17 |
| SP4 | GGE | 234 | 9.99E-01 | 3.72E-02 | 3.74E-01 | 9.27E-01 | 1.72E-04 | 7.11E-01 | 3.59E-02 | 4.25E-13 |
| SCN2A | DEE | 410 | 3.38E-01 | 5.84E-01 | 2.92E-13 | 6.28E-08 | 1.06E-04 | 4.16E-04 | 2.63E-02 | 3.40E-12 |
| STXBP1 | DEE | 56 | 6.43E-01 | 3.12E-01 | 5.48E-09 | 8.89E-12 | 1.87E-02 | 8.43E-05 | 2.22E-05 | 6.54E-11 |
| SLC6A1 | DEE | 60 | 2.71E-01 | 2.38E-01 | 2.69E-11 | 3.82E-07 | 2.64E-01 | 1.71E-04 | 1.82E-03 | 6.93E-11 |
| KCNQ2 | DEE | 149 | 1.18E-02 | 2.03E-01 | 5.83E-07 | 2.63E-01 | 2.60E-02 | 3.44E-02 | 2.75E-01 | 2.51E-09 |
| KCNB1 | DEE | 121 | 6.65E-02 | 2.87E-01 | 2.01E-04 | 6.63E-09 | 8.91E-01 | 1.32E-03 | 4.61E-04 | 9.46E-09 |
| RYR2 | GGE | 720 | 6.86E-01 | 2.36E-02 | 1.27E-01 | 2.03E-15 | 3.22E-01 | 6.45E-01 | 8.60E-06 | 5.17E-08 |
| SATB1 | DEE | 84 | 1.23E-01 | 4.81E-03 | 2.19E-01 | 9.31E-14 | 5.35E-02 | 4.29E-01 | 4.46E-02 | 5.28E-08 |
| CACNA1A | DEE | 278 | 1.74E-01 | 7.73E-01 | 1.55E-07 | 5.27E-04 | 3.97E-01 | 1.31E-02 | 3.67E-01 | 7.86E-08 |
| DYRK1A | DEE | 95 | 9.56E-01 | 8.94E-01 | 5.87E-01 | 2.01E-13 | 1.51E-01 | 7.16E-01 | 3.49E-04 | 2.52E-07 |
| CACNA1A** | GGE | 289 | 2.02E-04 | 8.14E-01 | 5.88E-02 | 4.10E-05 | 1.84E-01 | 1.48E-01 | 7.81E-03 | 5.82E-07 |
| ATP2B2** | DEE | 191 | 4.69E-04 | 6.40E-01 | 7.60E-01 | 5.93E-08 | 3.70E-01 | 2.03E-01 | 6.22E-03 | 2.74E-06 |
| CACNA1B** | GGE | 370 | 1.14E-01 | 9.62E-01 | 2.14E-02 | 7.80E-08 | 5.50E-02 | 2.14E-02 | 2.48E-04 | 5.12E-06 |
| KDM5B** | DEE | 268 | 7.38E-01 | 4.12E-01 | 6.16E-03 | 1.76E-17 | 2.87E-07 | 7.29E-01 | 2.71E-03 | 3.10E-04 |

We will first discuss the genes meeting the latter “importance” criterion, designated by the pink-highlighted rows in Table 1: *KCNQ2* (DEE) [MIM: 602235], *RYR2* (GGE) [MIM: 180902], *CACNA1A* (GGE) [MIM: 601011], *ATP2B2* (DEE) [MIM: 108733], *CACNA1B* (GGE) [MIM: 601012], and *KDM5B* (DEE) [MIM: 605393]. These genes can be partitioned into those that have established roles in epilepsy presentation and those genes whose role in epilepsy was inconclusive prior to this study. Among the established genes, *KCNQ2, CACNA1A*, and *CACNA1B* each encode ion-channel subunits essential for neuronal excitability and have long been implicated in epilepsy pathophysiology. Mutations in *KCNQ2* are associated with a spectrum of neonatal-onset epilepsy syndromes, including benign familial neonatal seizures (BFNS [MIM: 121200]) and developmental and epileptic encephalopathy (DEE) (https://www.kcnq2cure.org/kcnq2-epilepsy). Likewise, *CACNA1A* is linked to various types of epilepsy, including DEE (https://www.cacna1a.org/what-is-cacna1a), and mutations in *CACNA1B* have been associated with disorders ranging from rare episodic ataxia [MIM: 108500] to genetic generalized epilepsy (GGE).^22^,23 Within the unified model, *KCNQ2* reaches an overall p-value of 2. 51 × 10^−9^, driven primarily by constraint and pathogenicity effects (p-values of 1. 18 × 10^−2^ and ^−7^, respectively). Genes *CACNA1A* and *CACNA1B* show unified p-values of 5. 82 × 10^−7^and 5. 12 × 10^−6^, respectively, with both signals dominated by pLoF effects (4. 10 × 10^−5^ and 7. 80 × 10^−8^).

Several additional genes demonstrate newly significant or substantially strengthened associations under the unified framework. ATP2B2, which encodes a plasma-membrane calcium-transporting ATPase, achieves an overall p-value of 2. 74 × 10^−6^, improving upon its previously nonsignificant Epi25 result. Its signal is driven by pLoF effects (5. 93 × 10^−8^), consistent with prior evidence linking *ATP2B2* dysregulation to neuronal calcium signaling and seizure susceptibility.^24,25^ KDM5B, a chromatin-modifying gene associated with neurodevelopmental disorders,^26–28^ shows a unified p-value of 3. 10 × 10^−4^, dominated by a strong pLoF effect (*p* = 1. 76 × 10^−17^). Finally, RYR2, which encodes a ryanodine receptor critical for calcium-induced calcium release, displays a unified p-value of 5. 17 × 10^−8^ and a pronounced pLoF contribution (*p* = 2. 03 × 10^−16^), improving substantially over the Epi25 PTV analysis (*p* = 8. 60 × 10^−6^). Two recent studies have identified *RYR2* mutations in children with benign epilepsy of childhood with centrotemporal spikes (BECTS [MIM: 117100]) and focal epilepsy [MIM: 604364].^29^,30 Together, these results highlight that incorporating constraint-based, structural, and functional annotations into a single meta-regression substantially increases sensitivity to detect both known and emerging epilepsy genes while maintaining biological interpretability across distinct variant classes and epilepsy phenotypes.

Now, we consider the genes not meeting the “importance” criterion that meet the exome-wide significance threshold. Three main patterns emerge.

1. **Established DEE/GGE genes whose signals strengthen under integration**. A couple ion-channel and synaptic genes already discussed above—*KCNQ2, CACNA1A and RYR2*—now exceed the exome-wide significance threshold despite sub-threshold Epi25 results. These improvements are driven by moderators consistent with known biology (for example, pLoF effects for calcium-handling genes and constraint or pathogenicity effects for *KCNQ2*), so their mechanisms are not restated here.
2. **Synaptic release, scaffold, and inhibitory-tone genes newly cross the threshold**. *STXBP1* (DEE) [MIM: 602926] and *SLC6A1* (DEE) [MIM: 137165] show strong moderator support from pLoF and pathogenicity terms that elevate their overall significance beyond what Epi25 reported for damaging missense or truncating variants. Similarly, *DLG4* (DEE) [MIM: 602887] and *KCNB1* (DEE) [MIM: 600397] achieve exome-wide significance through coherent constraint and pLoF contributions, reinforcing the role of synaptic and potassium-channel pathways in epileptic encephalopathy.
3. **Regulatory and emerging candidates**. The unified model also brings to significance several transcriptional or chromatin regulators—*SETD1B* (GGE) [MIM: 611055], *SATB1* (DEE) [MIM: 602075], and *SP4* (GGE) [MIM: 600540]—whose strongest moderators are constraint and missense pathogenicity. Finally, Table 1 identifies more exploratory candidates such as *PCDHAC2* (DEE) [MIM: 606321], which is mechanistically plausible given its role in neuronal adhesion but currently lacks direct clinical validation.

Overall, Table 1 demonstrates that the unified framework systematically converts multiple near-miss Epi25 loci into exome-wide associations across distinct epilepsy subgroups while maintaining biological interpretability by variant class. The highly-significant rows of Table 1 isolates those genes whose associations now cross the study-wide boundary, underscoring how the integrated model enhances detection power for both established and emerging epilepsy genes. We visually summarize these results across the four represented gene groups in Figure 5.

**Figure 5:**
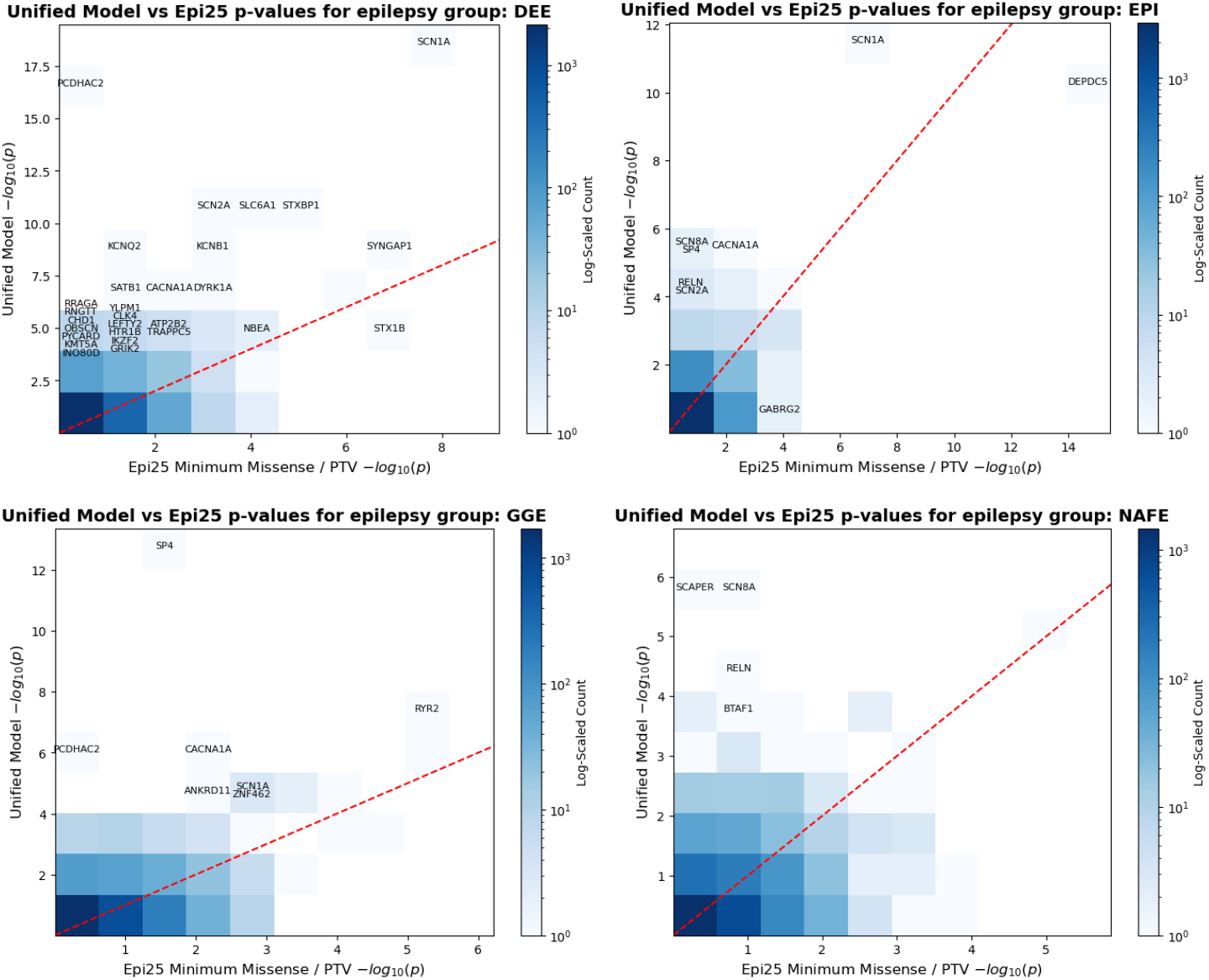
Comparison of unified model p-values and minimum Epi25 damaging missense or PTV p-values across four epilepsy gene groups. Each subplot shows a 2D histogram (log-scaled count) of − *log*_10_(*p*) values. The red dashed diagonal indicates equivalence between the two models. Gene names are annotated where the absolute *log*_10_(*p*) difference exceeds 2 and at least one model achieves *p* < 10^−4^; labels were grouped by bin coordinates to improve readability and vertically stacked to avoid overlap.

Figure 5 summarizes the comparative performance of the unified meta-regression model relative to the Epi25 burden analyses across four epilepsy subgroups: developmental and epileptic encephalopathy (DEE), generalized genetic epilepsy (GGE), focal epilepsy (EPI), and non-acquired focal epilepsy (NAFE). Each panel plots the unified model p-values against the minimum of the Epi25 missense or protein-truncating variant (PTV) p-values for the same gene, with the red diagonal indicating equivalence between the two approaches. Points below the diagonal represent genes for which the unified model yields stronger associations. Across all four epilepsy subgroups, the majority of genes fall below this line, illustrating that the unified model systematically improves gene-level significance relative to the Epi25 analysis. This trend is especially pronounced for genes with intermediate Epi25 signals—those near but not surpassing exome-wide significance—where the unified framework integrates complementary sources of evidence (constraint, pathogenicity, and pLoF) to amplify biologically meaningful signals. Although many canonical epilepsy genes—including *SCN1A* [MIM: 182389], *SCN2A* [MIM: 182390], and *STXBP1* [MIM: 602926] in DEE and *RYR2* and *CACNA1A* in GGE—exhibit stronger signals under the unified framework, a subset—notably STX1B [MIM: 601485] in DEE and DEPDC5 [MIM: 614191] and GABRG2 [MIM: 147164] in EPI—show comparatively weaker associations. This discrepancy likely reflects the weighting of annotation priors in the meta-regression, where genes with higher predicted tolerance or limited AlphaMissense or pLoF support receive less weight despite burden enrichment. Such cases may therefore represent genes where statistical burden and biological annotation provide complementary but non-concordant evidence. Overall, these results demonstrate that the unified approach both recapitulates known epilepsy genes and enhances statistical power to detect moderate-effect associations across heterogeneous epilepsy phenotypes.

### Schizophrenia

The focus of this paper is on the application of our unified meta-regression model to epilepsy; however, there are likely extensions of this methodology to other neurological and psychiatric disorders. In this brief subsection, we discuss preliminary findings from extending the unified meta-regression framework to schizophrenia [MIM: SCZD] using data from the Schizophrenia Exome Sequencing Meta-analysis (SCHEMA) consortium.^31^ These findings, demonstrated below in Figure 6 and Table 2 are more mixed than our epilepsy findings, prompting more specific model development for each disease of interest.

**Table 2:** Comparison of p-values for each moderator of the unified model, the unified model p-value, and the SCHEMA p-value. Results are shown for the gene-group pairs with unified model p-value at most 1. 0 × 10^−4^, ordered in ascending order by unified model p-value.

| Gene Name | Group | # of Variants | Constraint p-value | GERP p-value | Pathogenicity p-value | pLoF p-value | Missense p-value | SCHEMA p-value | Unified Model p-value |
| --- | --- | --- | --- | --- | --- | --- | --- | --- | --- |
| PCDHGA5 | meta | 1460 | 4.55E-16 | 1.06E-01 | 1.11E-02 | 2.99E-01 | 7.85E-08 | 1.09E-01 | 5.64E-15 |
| XPO7 | meta | 241 | 5.28E-01 | 1.14E-01 | 6.80E-03 | 1.12E-09 | 3.54E-04 | 7.18E-09 | 6.78E-08 |
| SETD1A | meta | 627 | 1.77E-01 | 9.74E-01 | 4.98E-02 | 4.38E-09 | 2.62E-02 | 2.00E-12 | 1.10E-06 |
| KLRC2 | meta | 50 | 1.78E-06 | 6.19E-04 | 2.29E-02 | 8.60E-01 | 6.50E-01 | 3.42E-01 | 3.59E-06 |
| ATP9A | meta | 278 | 8.71E-01 | 8.49E-01 | 4.40E-01 | 2.54E-11 | 4.46E-03 | 3.72E-04 | 3.59E-06 |
| ASH1L | meta | 918 | 2.63E-02 | 6.04E-01 | 5.55E-01 | 9.05E-01 | 1.25E-01 | 3.77E-05 | 6.32E-06 |
| DNAI7 | meta | 202 | 8.82E-01 | 8.55E-03 | 2.82E-02 | 8.46E-01 | 2.50E-01 | 1.51E-02 | 7.87E-06 |
| TLK2 | meta | 111 | 5.95E-01 | 3.42E-03 | 1.76E-05 | 6.52E-04 | 1.53E-01 | 3.45E-03 | 1.06E-05 |
| GRIN2A | meta | 467 | 3.13E-01 | 9.05E-01 | 4.51E-02 | 4.40E-08 | 7.53E-03 | 7.37E-07 | 1.51E-05 |
| AKAP11 | meta | 667 | 9.94E-01 | 8.49E-01 | 6.31E-01 | 3.62E-11 | 6.17E-02 | 8.28E-06 | 2.29E-05 |
| PCDHA4 | meta | 1027 | 1.64E-01 | 3.18E-06 | 1.11E-02 | 3.87E-01 | 6.52E-01 | 4.34E-01 | 4.61E-05 |
| SCN2A | meta | 689 | 7.60E-01 | 7.44E-01 | 5.32E-06 | 1.80E-02 | 3.88E-01 | 2.32E-02 | 5.48E-05 |
| SRRM2 | meta | 97 | 6.37E-01 | 9.08E-01 | 8.73E-11 | 8.90E-11 | 5.07E-02 | 1.53E-05 | 5.49E-05 |
| UTP20 | meta | 959 | 5.63E-05 | 3.57E-01 | 4.49E-01 | 1.11E-04 | 5.11E-02 | 1.98E-01 | 9.41E-05 |

**Figure 6:**
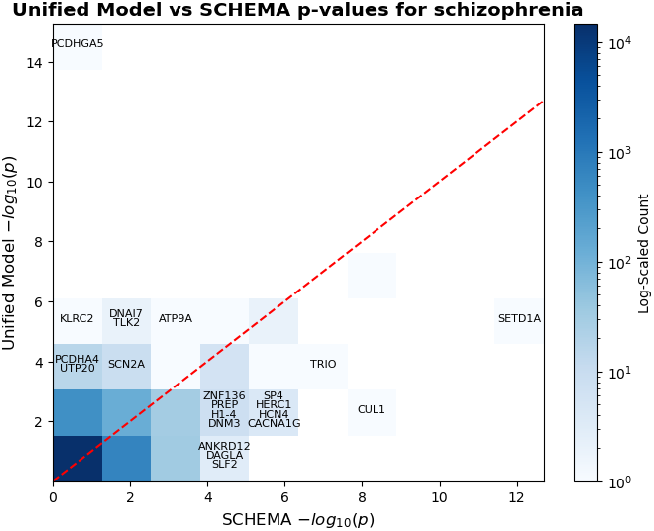
Comparison of unified model p-values and SCHEMA p-values across genes in the schizophrenia “meta” group. The plot shows a 2D histogram (log-scaled count) of − *log*_10_(*p*) values. The red dashed diagonal indicates equivalence between the two models. Gene names are annotated where the absolute *log*_10_(*p*) difference exceeds 2 and at least one model achieves *p* < 10^−4^; labels were grouped by bin coordinates to improve readability and vertically stacked to avoid overlap.

The SCHEMA dataset represents one of the largest and most comprehensive exome-sequencing resources for psychiatric genetics, aggregating exome data from 24,248 schizophrenia cases, 97,322 controls, and 3,402 parent–proband trios across multiple international cohorts. We use the SCHEMA Phase I dataset as a benchmark to evaluate whether our unified meta-regression can replicate or refine these associations by integrating complementary information from sequence-based constraint, structural pathogenicity predictions, and predicted loss-of-function effects. Unlike the epilepsy analyses, where phenotype subgroups are clearly defined, the schizophrenia dataset aggregates all cases under a single “meta” group, allowing us to assess the generalizability of our framework across a more heterogeneous, polygenic psychiatric disorder.

Building on this benchmark, we next compare the unified model p-values with those from the SCHEMA meta-analysis to assess whether our integrative approach captures known risk genes or identifies additional candidates with similar biological signatures. Ideally, the unified p-values should equal or surpass the strength of the SCHEMA benchmark, indicating comparable or improved performance. However, results were mixed. The largest improvement was observed for *PCDHGA5*, which displayed a marked gain in significance under the unified framework (*p* = 5. 64 × 10^−15^ versus *p* = 1. 09 × 10^−1^ in SCHEMA). Moderate improvements were also seen for *KLRC2* [MIM:602891], *DNAI7* [MIM: 616906], and *TLK2* [MIM: 608439]. In contrast, several genes that are highly significant in SCHEMA, including *SETD1A* [MIM: 611052], *CUL1* [MIM:603134], and *TRIO* [MIM: 601893], did not replicate under the unified model, reflecting incomplete power and potential model mismatches between epilepsy and schizophrenia. Figure 6 summarizes these results across all genes analyzed in the SCHEMA dataset, comparing the unified model and SCHEMA p-values by gene.

Overall, these results indicate that while the unified model can recover and occasionally strengthen individual schizophrenia associations, its performance is less consistent than in epilepsy. This limitation likely stems from smaller effect sizes, higher genetic heterogeneity, and the absence of explicit modeling of *de novo* mutations—an important contributor to rare variant risk in schizophrenia as demonstrated by the SCHEMA consortium. Future work should therefore prioritize integrating de novo mutation information and increasing sample size to improve the power and stability of unified model estimates in schizophrenia and other highly polygenic psychiatric disorders.

## Discussion

Our study introduces a unified statistical framework that integrates sequence-level constraint, structure-based missense pathogenicity, predicted loss-of-function (pLoF) annotations, and rejected substitution (RS) scores to improve detection of epilepsy-associated genes. By combining Hidden Markov Model (HMM)–derived constraint predictions with AlphaMissense (AM) pathogenicity probabilities, pLoF/missense indicators, and GERP RS scores within a weighted meta-regression model, we demonstrate that distinct biological signals can be systematically combined to enhance statistical power in rare variant association studies. The unified model consistently identifies both known epilepsy genes and novel candidates that fall below genome-wide significance in standard population-based analyses, illustrating how sequence- and structure-informed priors can refine gene discovery in heterogeneous neurodevelopmental disorders.

Our findings emphasize the complementary roles of constraint, pathogenicity, and functional loss in capturing diverse aspects of mutational intolerance. Genes such as *KCNQ2, CACNA1A*, and *CACNA1B*—each encoding ion-channel subunits essential for neuronal excitability—show improved gene-level significance driven by different model components: constraint and pathogenicity for *KCNQ2*, and pLoF for *CACNA1A* and *CACNA1B*. Beyond these well-established genes, the unified model strengthens signals for genes with limited prior evidence of causality, including *ATP2B2, KDM5B*, and *RYR2*. The discovery of significant or suggestive associations in these genes, each linked to calcium signaling, chromatin regulation, or neuronal excitability, highlights the model’s ability to detect biologically plausible yet previously underpowered relationships. Together, these results underscore the value of integrating orthogonal predictors of mutational impact into a single inferential framework for gene-level discovery.

While promising, the utility of this model is bounded by the accuracy and completeness of its inputs. Structural pathogenicity scores such as AM rely heavily on protein coverage and model quality, whereas constraint inference is sensitive to sequencing depth and variant ascertainment. As future datasets incorporate more diverse ancestries and deeper coverage, the constraint model can be retrained to reduce bias and improve generalization. Methodologically, extending the HMM into a higher-order or neural-sequence model—such as a recurrent or transformer-based architecture—could capture more complex dependencies in mutational depletion patterns. Similarly, nonlinear extensions of the meta-regression could allow for interactions among predictors (e.g., between structural intolerance and constraint) that remain unexplored in the current linear framework. Moreover, because the framework emphasizes depletion-based and loss-of-function–oriented predictors, it may underrepresent gain-of-function associations—an important consideration for epilepsy genes such as SCN2A and KCNQ2, where different mutational mechanisms can produce distinct phenotypes.^32,33^ Future extensions could integrate functional assay data or gain-of-function–specific pathogenicity predictors to better capture this class of effects.

Finally, this unified framework has broader applicability beyond epilepsy. Because it models joint variant-level contributions from multiple biological priors, the approach can be extended to other neurodevelopmental and psychiatric disorders—such as autism, schizophrenia, and bipolar disorder—where rare variant heterogeneity limits discovery power. In this study, we applied the model to an independent schizophrenia dataset from the SCHEMA consortium as a proof of generalizability. The unified model recovered several known risk loci and strengthened associations for select candidates (for example, *PCDHGA5* and *TLK2*), though its overall performance was less consistent than in epilepsy, likely reflecting smaller effect sizes and greater locus heterogeneity in schizophrenia. This cross-disorder validation highlights both the adaptability of the framework and its dependence on disease architecture, supporting future applications to other large-scale sequencing studies aimed at integrating functional priors into rare variant discovery.

## Supporting information

Full meta-regression results for epilepsy

## Data Availability

All data produced are either available on github or upon reasonable request to the authors

https://github.com/healthcare-medicine-ai/wgs-constraint-llm

## Supporting Information

### Data and Code Availability

The code for our model development and data analysis is hosted on a GitHub repository at https://github.com/healthcare-medicine-ai/wgs-constraint-llm, along with most data and results. The full WES constraint predictions generated for AoU and RGC-ME are available at https://doi.org/10.6084/m9.figshare.27184245.v1.

## Acknowledgements

M.A.R. is in part supported by National Human Genome Research Institute (NHGRI) under award R01HG010140, and by the National Institutes of Mental Health (NIMH) under award R01MH124244 both of the National Institutes of Health (NIH).

Some of the computing for this project was performed on the Sherlock cluster. We would like to thank Stanford University and the Stanford Research Computing Center for providing computational resources and support that contributed to these research results. The content is solely the responsibility of the authors and does not necessarily represent the official views of the funding agencies; funders had no role in study design, data collection and analysis, decision to publish, or preparation of the manuscript.

## Declaration of interests

M.A.R. is a cofounder of Broadwing Bio and a consultant for Insitro.

Declaration of generative AI and AI-assisted technologies in the writing process

During the preparation of this work the authors used perplexity.ai and chat-gpt in order to investigate the relationship between epilepsy and certain genes in the existing literature and synthesize references to related work. After using this tool, the authors reviewed and edited the content as needed and take full responsibility for the content of the publication.

## Online resources

Regeneron Genetics Center Million Exome Variant Browser https://rgc-research.regeneron.com/me/license-and-terms-of-use

AlphaMissense predictions https://console.cloud.google.com/storage/browser/dm_alphamissense

GERP RS Scores

https://genome-asia.ucsc.edu/cgi-bin/hgTables?db=hg19&hgta_group=compGeno&hgta_track=allHg19RS_BW&hgta_table=allHg19RS_BW&hgta_doSchema=describe+table+schema

gnomAD v4.1.0 https://gnomad.broadinstitute.org/

Epi25 Browser https://epi25.broadinstitute.org/

SCHEMA Browser https://schema.broadinstitute.org/

## Supplementary Materials

**Table S1:**
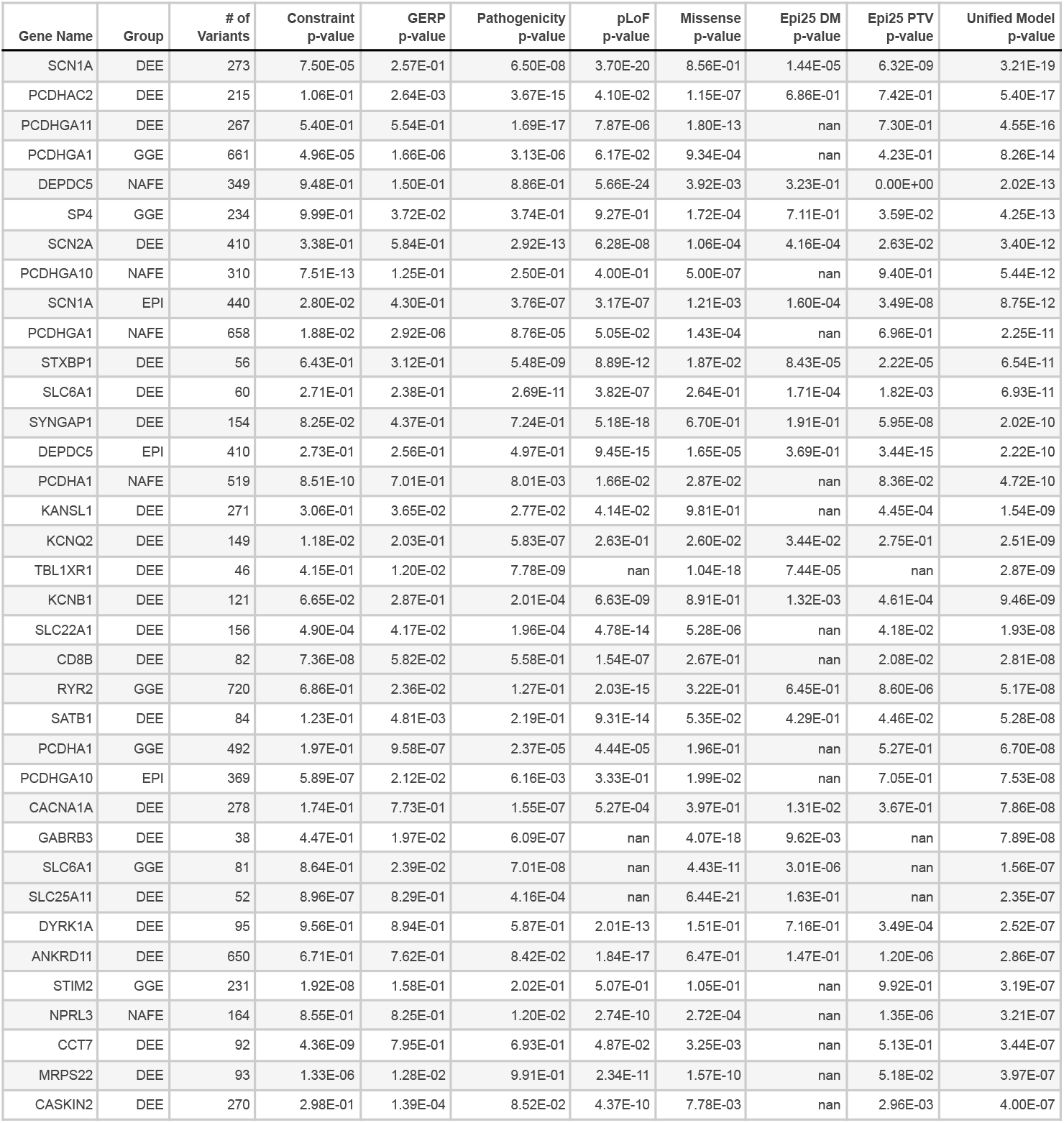
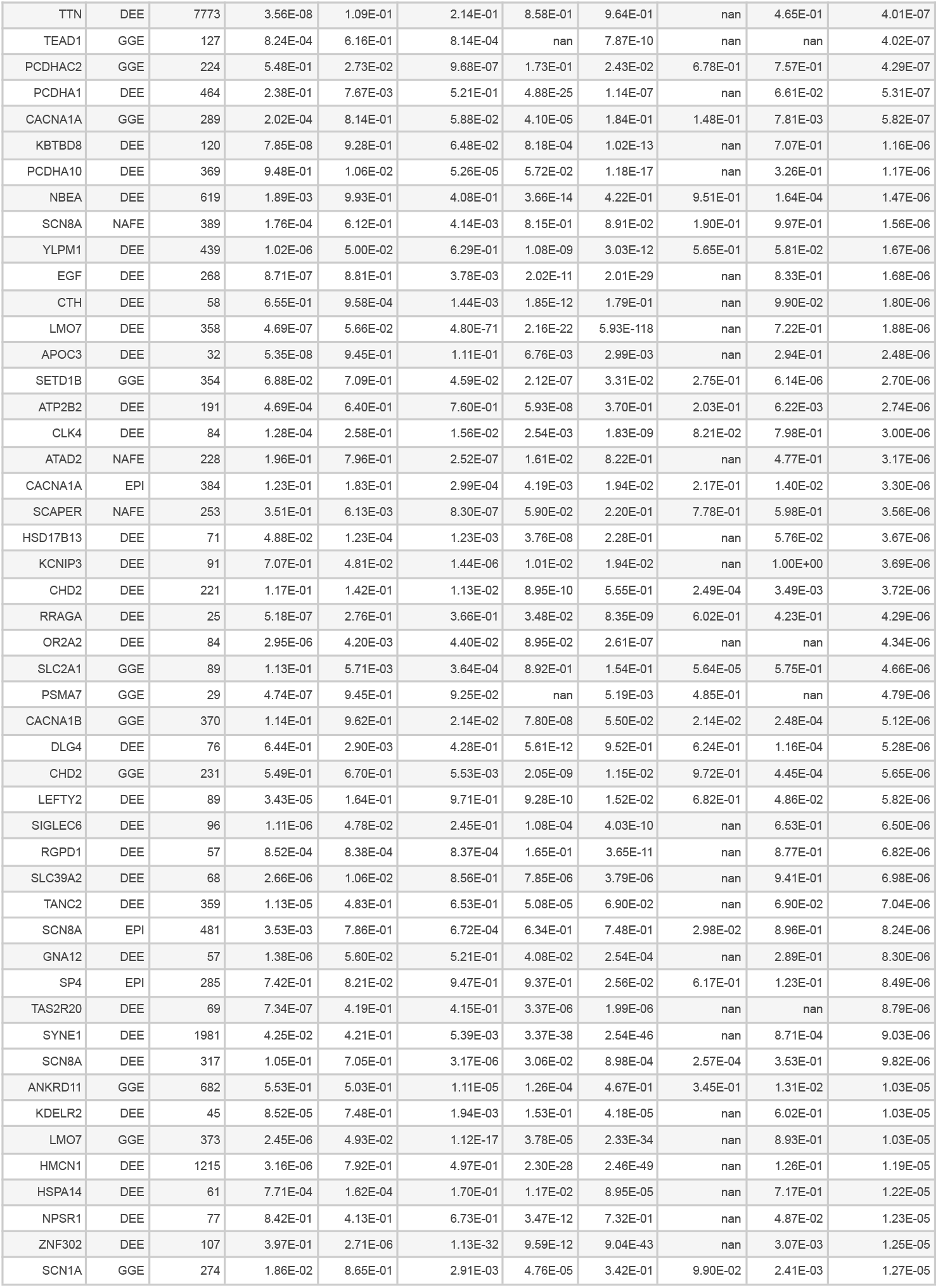
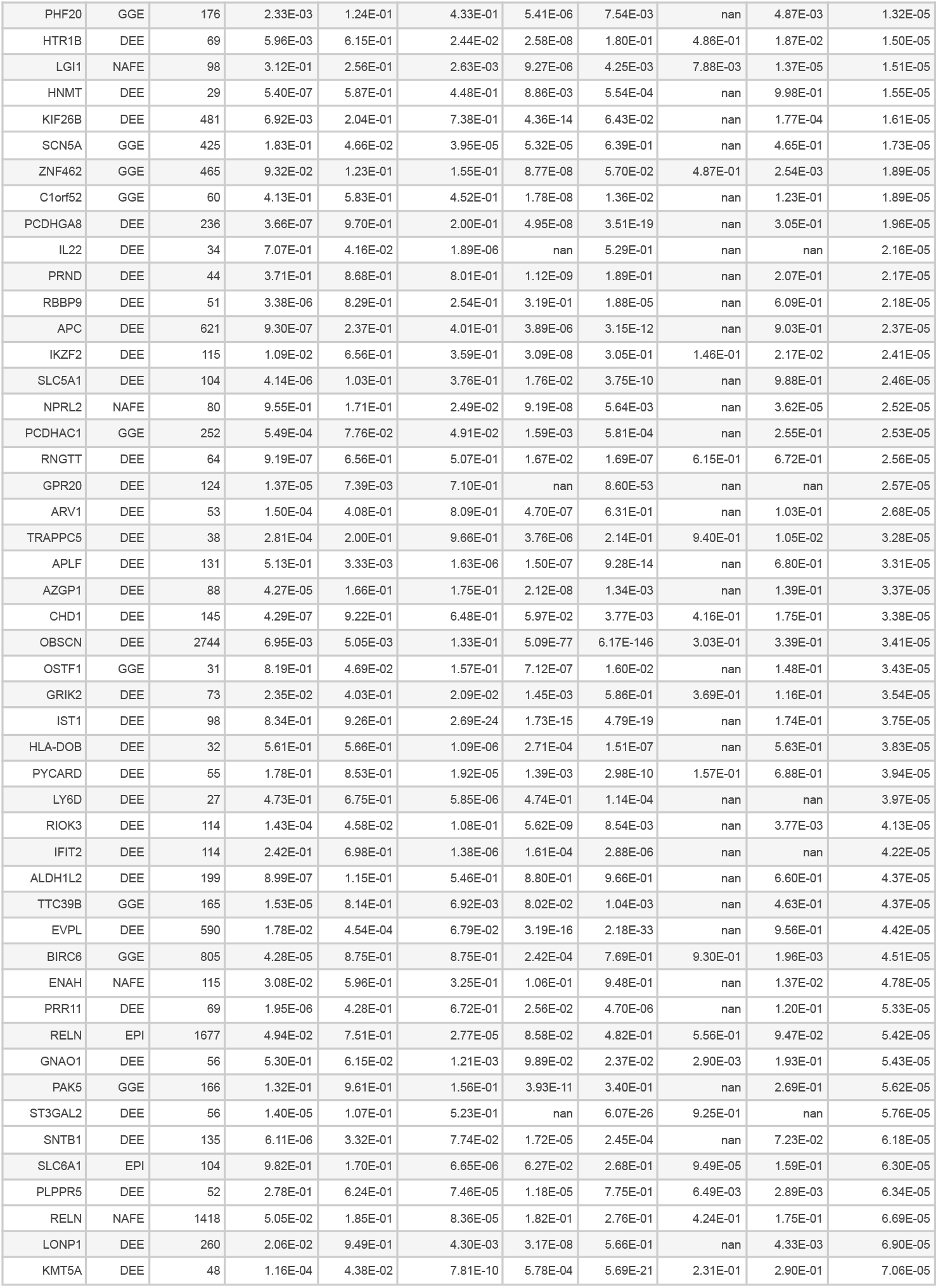
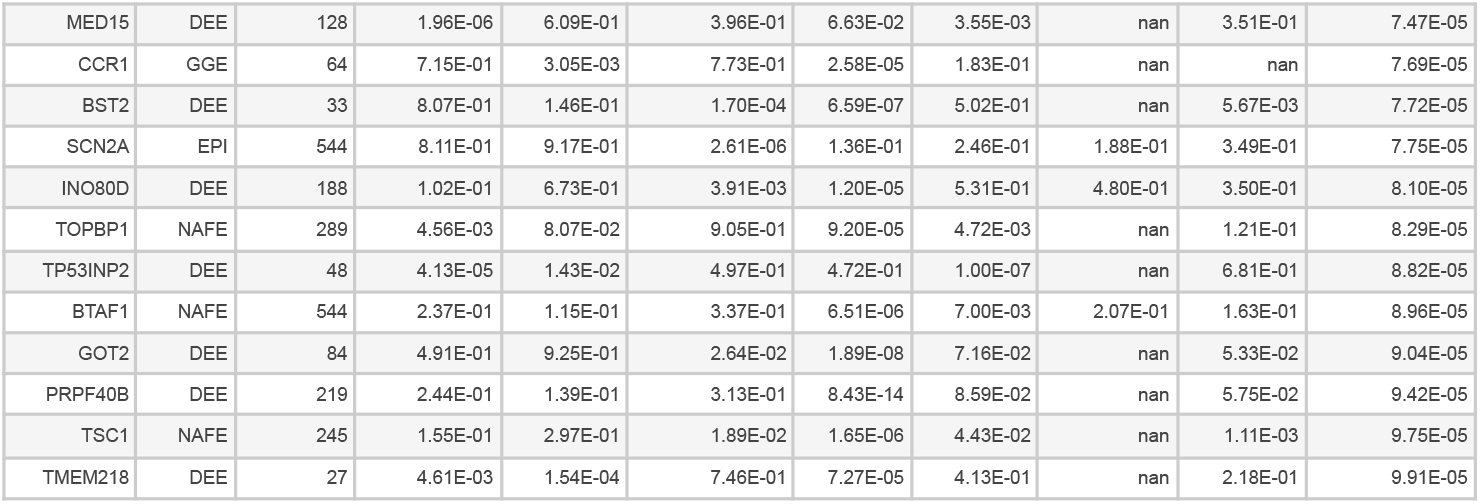
Comparison of p-values for each moderator of the unified model, the unified model p-value, and the Epi25 damaging missense and PTV p-values across four epilepsy gene groups. Results are shown for the gene-group pairs with unified model p-value < 1e-4, ordered by unified model p-value.

